# Diagnostic comparison of three fully automated chemiluminescent immunoassay platforms for the detection of SARS-CoV-2 antibodies

**DOI:** 10.1101/2020.10.07.20207696

**Authors:** Debaprasad Parai, Girish Chandra Dash, Hari Ram Choudhary, Annalisha Peter, Usha Kiran Rout, Rashmi Ranjan Nanda, Jaya Singh Kshatri, Srikanta Kanungo, Subrata Kumar Palo, Jyotirmayee Turuk, Sanghamitra Pati, Debdutta Bhattacharya

## Abstract

The whole world is battling against coronavirus disease-19 (COVID-19) pandemic caused by severe acute respiratory syndrome coronavirus 2 (SARS-CoV-2). Various strategies are taken to curb the spread of the virus and to move out from the enforced lockdown stage. Serological tests are the neediest diagnostic and surveillance tool to complement the gold standard molecular diagnostic method to track down the transmission rate of SARS-CoV-2. Automated chemiluminescent immunoassay (CLIA) based analyzers become highly demanding platforms both to clinicians and policy makers for the detection anti-SARS-CoV-2 antibodies. In this study, serum from 594 patients positive for COVID-19 and 100 samples from pre-COVID cases were tested by three automated platforms: Abbott architect i2000SR, Roche cobas e411 and Yhlo iFlash 1800 and their diagnostic accuracy were compared. All three platforms showed high specificity as claimed by manufacturer. Clinical sensitivities of the machines were calculated as 64.48% (58.67-70.3) for Abbott, 80.48% (76.62-84.34) for Roche and 76.94% (72.65-81.23) for Yhlo. The Cohen’s kappa value was determined from 0.69-0.89 when inter-rater agreements were calculated. The area under the curves (AUC) values demonstrated Roche Cobas e411 as the diagnostically most accurate platform among the three CLIA analyzers.

## Introduction

The world is still dealing with the coronavirus disease-19 (COVID-19) pandemic caused by severe acute respiratory syndrome coronavirus 2 (SARS-CoV-2) since it started in December 2019 (1,2). Accurate and speedy diagnosis of SARS-CoV-2 infection is very much needed for prompt and effective patient care. Nasopharyngeal swab (NPS) followed by reverse-transcriptase polymerase chain reaction (RT-qPCR) is the gold standard of molecular diagnosis for SARS-CoV-2 detection (3). However, sometimes it fails to demonstrate the complete picture of the rapid transmission of the virus through communities (4). Hence, serological test is believed to be another important diagnostic tool along with swab test. This test is also called as antibody test as it detects anti-SARS-CoV-2 immunoglobulins which are usually formed in patient body as early as by 1 week and in general within 2-3 weeks from the infection onset (5,6). Antibody tests are a useful surveillance tool to track down the prevalence of COVID-19 epidemiology and to assess the current immune status of a certain community. Serological tests are also useful for the policy makers to decide the lockdown entry and exit strategies, particularly in this second wave of pandemic (7-9).

Currently, there are several serological assays are available in the market to detect anti-SARS-CoV-2. These are mainly relied on the principle of quantitative laboratory-based enzyme linked immunosorbent assay (ELISA), chemiluminescent immunoassay (CLIA), or a qualitative point-of-care test (POCT). Recently developed fully automated analysers based on CLIA technology are having high potential with large throughput (10,11). However, to detect the accurate one is of great challenge. In this study, we compared three such automated analysers: ARCHITECT i2000SR (Abbott Laboratories, Chicago, USA), Cobas e411 (Roche Diagnostics GmbH, Mannheim, Germany) and iFlash 1800 (Shenzhen Yhlo Biotech Co. Ltd., Shenzhen, China) to identify their diagnostic accuracy.

## Methods

### Collection of serum sample

Serum samples were collected from recovered COVID-19 patients after 4 weeks and not more than 8 weeks from the detection of SARS-CoV-2 infection. All the COVID-19 patients were confirmed positive to SARS-CoV-2 tested by oropharyngeal and nasopharyngeal swabs followed by RT-PCR. COVID-19 patients were informed about the serological test with proper written consent.

A total of 594 subjects were chosen for this study between 23^th^ July 2020 and 14^th^ September 2020. The 100 samples collected during pre COVID period (August, 2019) and stored in RMRC-Bhubaneswar sample repository were used as control. The study was approved by Institutional Ethics Committee.

### Test method

All three different automated machines can qualitatively detect anti-SARS-CoV-2 antibodies based on the two-step immunoassay principle. Abbott made ARCHITECT i2000SR platform uses chemiluminescent microparticle immunoassay (CMIA) technology for the detection of immunoglobulin class G (IgG) antibodies against the nucleocapsid protein of SARS-CoV-2 from human serum. The specificity of SARS-CoV-2 IgG assay in this platform was 99.63% (95% confidence of interval [CI]: 99.05-99.90%) and sensitivity was 100% (95% CI: 95.89-100%) when tested after 14 days post-symptom onset as per the manufacturer. The cut-off value was 1.4 index. Second automated machine Cobas e411 by Roche Diagnostics determined the presence of antibodies including IgG against SARS-Cov-2 based on a patented electro chemiluminescence (ECL) technology. The test principle was a sandwich assay using biotinylated SARS⍰CoV⍰2⍰specific recombinant nucleocapsid protein and streptavidin-coated microparticles. The specificity of Elecsys Anti-SARS-CoV-2 was reported as 99.81 % (95% CI: 99.65-99.91%) and the sensitivity was 100 % (95% CI: 88.1-100%) as per manufacturer when the serum was tested after 14 days post SARS-Cov-2 confirmation. The cut-off value of this assay was 1.0 COI. The third one, Yhlo Biotech manufactured iFlash-SARS-CoV-2 IgG was a paramagnetic particle based chemiluminescent immunoassay (CLIA) to determine the IgG antibodies against SARS-CoV-2 nucleocapsid and spike protein. According to the manufacturer, the clinical specificity and sensitivity of this assay were 96.3% and 97.3%, respectively. Result was interpreted either as non-reactive: < 10.0 AU/mL or reactive: > 10.0 AU/mL.

### Statistical analysis

Descriptive statistical analyses were performed by SPSS software (IBM SPSS statistics for Windows, version 24.0, Armonk, NY). The agreement among the automated platforms were measured by Cohen’s kappa (k). Specificity, sensitivity, negative predictive value (NPV) and positive predictive value (PPV) were calculated for each individual assay. Area under the curves (AUC) were compared from the receiver operator characteristic (ROC) curves of the three different platforms. *p* value <0.05 was considered as statistically significant.

## Results

A total 594 samples from recovered COVID-19 patients and 100 pre-COVID serum samples were analysed in the 3 CLIA platforms. Table 1 describes the calculated specificity and sensitivity of all three automated SARS-CoV-2 antibody detecting platforms. Abbott showed a specificity of 99% (95% CI: 97.09-100.92%) and with sensitivity of 64.48% (95% CI: 58.67-70.3). The PPV and NPV were determined as 99.74% (99.25-100.24) and 31.94% (16.1-47.78), respectively. Specificity for iFlash-SARS-CoV-2 IgG assay was 100%, whereas sensitivity was calculated as 76.94% (72.65-81.23). This platform had a PPV of 100% and NPV of 42.2 (27.67-56.72). Roche Elecsys Anti-SARS-CoV-2 insert recorded highest sensitivity 80.48% (76.62-84.34) compared to other two chemiluminescent platforms and the specificity (100%) was same with Yhlo machine. The PPV and NPV of Roche analyser were 100% (100.0-100.0) and 46.3% (32.3-60.3), respectively.

**Table 1.** Analytical specificities, sensitivities, positive predictive values (PPV) and negative predictive values (NPV) with 95% confidence intervals (CI) for SARS-CoV-2 antibody.

|  | **Abbott** | **Roche** | **Yhlo** |
| --- | --- | --- | --- |
| **Insert name** | SARS-CoV-2 IgG | Elecsys Anti-SARS-CoV-2 | iFlash- SARS-CoV-2 IgG |
| **Cut-off** | 1.0 Col | 1.4 Index | 10.0 AU/mL |
| **SARS-CoV-2 positive sera** | 383/594 | 478/594 | 457/594 |
| **Control negative sera** | 99/100 | 100/100 | 100/100 |
| **Specificity (95% CI)** | 99.0% (97.09-100.92) | 100% (100.0-100.0) | 100% (100.0-100.0) |
| **Sensitivity (95% CI)** | 64.48% (58.67-70.3) | 80.48% (76.62-84.34) | 76.94% (72.65-81.23) |
| **PPV** | 99.74 (99.25-100.24) | 100 (100.0-100.0) | 100 (100.0-100.0) |
| **NPV** | 31.94 (16.1-47.78) | 46.3 (32.3-60.3) | 42.2 (27.67-56.72) |

The ROC performance curves showed that Roche platform had the highest AUC value of 0.929 (95% CI: 0.910 – 0.948). Architect i2000SR gave AUC value of 0.863 (95% CI: 0.836 - 0.889) and for iFlash 1800, it was calculated as 0.897 (95% CI: 0.875 – 0.920).

Inter-rater agreement between Roche and Abbott was measured by k value of 0.694 (95% CI: 0.641 - 0.746) which is a good agreement (Table 2). The percent agreement between these two platforms was 85.3%. The k value was determined as 0.758 (95% CI: 0.709 - 0.807) between Yhlo vs Abbott analyzers and 88.3% was the percent agreement. A very good agreement (k =0.892; 95% CI: 0.856 - 0.927) was found between Elecsys Anti-SARS-CoV-2 vs iFlash-SARS-CoV-2 IgG. The percent agreement of 95.2% demonstrated the best inter-rater reliability among the raters (table 2).

**Table 2.** Inter-rater agreement (Cohen’s kappa; k) between three automated platforms for SARS-CoV-2 antibodies. Value of k <0.20 poor agreement, 0.21-0.40 fair agreement, 0.41- 0.60 moderate agreement, 0.61-0.80 good agreement, and 0.81-1.00 very good agreement.

| Roche vs Abbott |  | Abbott |  | Total |
| --- | --- | --- | --- | --- |
|  |  | Positive | Negative |  |
| Roche | Positive | 380 (54.8%) | 98 (14.1%) | 478 (68.9%) |
|  | Negative | 4 (0.6%) | 212 (30.5%) | 116 (31.1%) |
| Total |  | 384 (55.3%) | 310 (44.7%) | 694 (100%) |
| Kappa Value |  | 0.694 (0.641 - 0.746) |  |  |
| Agreement |  | 85.3% |  |  |

| Yhlo vs Abbott |  | Abbott |  | Total |
| --- | --- | --- | --- | --- |
|  |  | Positive | Negative |  |
| Yhlo | Positive | 380 (54.8%) | 77 (11.1%) | 457 (65.9%) |
|  | Negative | 4 (0.6%) | 233 (33.6%) | 137 (34.1%) |
| Total |  | 384 (55.3%) | 310 (44.7%) | 694 (100%) |
| Kappa Value |  | 0.758 (0.709 - 0.807) |  |  |
| Agreement |  | 88.3% |  |  |

| Roche vs Yhlo |  | Yhlo |  | Total |
| --- | --- | --- | --- | --- |
|  |  | Positive | Negative |  |
| Roche | Positive | 451 (65.9%) | 27 (3.9%) | 478 (68.9%) |
|  | Negative | 6 (0.9%) | 210 (30.3%) | 116 (31.1%) |
| Total |  | 457 (76.9%) | 137 (23.1%) | 694 (100%) |
| Kappa Value |  | 0.892 (0.856 - 0.927) |  |  |
| Agreement |  | 95.2% |  |  |

## Discussion

With the rapid spread of SARS-CoV-2 infection among the communities, rapid and bulk tests are urgently needed to determine the extent of COVID-19 pandemic at community level. Serological testing is a complementary test of conventional swab tests to predict the epidemiology prevalence. Serological surveys are useful tool to predict how far a community is from herd immunity (5,12). Routine antibodies tests are also necessary to identify the potential convalescent plasma donor for plasma therapy of critically ill COVID-19 patients (13). It is highly challenging to choose the best diagnostic platform for anti-SARS-CoV-2 antibodies test from the market available machines. To our best knowledge, the concordance of these three popular automated chemiluminescent assay platforms are evaluated for the first time in this study.

Currently, there is no gold standard for serological detection of anti-SARS-CoV-2 antibodies and comparative studies. Hence, we include a total of 100 pre-COVID samples as control set to determine the diagnostic accuracy. The chances to get anti-SARS-CoV-2 antibodies became higher after 14 days from the first detection. To test the diagnostic sensitivity, we included only those COVID-19 positive patients who had been confirmed at least 28 days before and recovered from the day of serum sample collection. Among the 594 positive sera samples, 378 samples gave positive or reactive whereas, 109 samples (18.35%) showed non-reactivity or negative in all three platforms. A 18.35% of non-reactive results revealed a majority of COVID-19 recovered patients were unable to produce detectable titre of antibodies. This result suggested us that the exact immunological response to SARS-CoV-2 infection is still poorly understood by the researchers. Both Roche and Yhlo platform were found to have 100% specificity, which is more than the manufacturer claimed value. In term of sensitivity, Roche insert showed highest sensitivity (80.48%) against anti-SARS-CoV-2 antibodies including IgG compared to both Abbott (64.48%) and Yhlo (76.94%). A study by Perkmann et al. showed a higher sensitivity for Abbott (84.6%) and Roche (89.2%) platforms although that might be because of the low number of recruited COVID-19 patients (n=65) (14). Similarly, another study with iFlash-SARS-CoV-2 IgG measured a sensitivity of 76.7% with 61 positive sera which is same (76.9%) with our result and thus corroborated our study with higher sample size (15).

The AUC values represented the diagnostic accuracy of among the three platforms and Roche gave the highest value of 0.929 at 95% CI. Inter-rater agreement those platforms were statistically good and found to be the highest between Roche and Yhlo. As per our study, Roche Cobas e411 automated chemiluminescent platform gave the best diagnostic accuracy. Statistically determined k value demonstrated other two machines were also not too behind from Roche as both of them showed good percent agreement. Elecsys Anti-SARS-CoV-2 assay is based on the nucleocapsid determination of SAR-CoV-2 and iFlash-SARS-CoV-2 IgG assay insert detected both nucleocapsid and spike proteins. However, both the assay platform showed an agreement of 95.2% which suggested that diagnostic accuracy was not truly depended on the target antigen as per this comparative study.

Since everyone around the world is looking forward to see the herd immunity in their respective localities and therefore the rapid serological testing is the high need of current time (16). The opportunity to test outside the laboratory is high to cover the larger population without putting the extra burden on the clinical laboratories and POCT may play an important role in doing so. However, mistakes in interpretation of test results, untrained staffs and low sensitivity & specificity could be the biggest roadblocks while performing the POCT as a tool for serological testing (17). Therefore, the automated machines are recommended for testing with high sensitivity and specificity to detect anti-SARS-CoV-2 antibodies and that is too in very short time.

In conclusion, detection of anti-SARS-CoV-2 antibodies are highly inconsistent throughout all the different automated chemiluminescent assay platforms. This study shows the diagnostics accuracy of three popular automated platform and compared their agreements when tested with higher sample size. This is the first such demonstration of these three platforms which would be very much helpful for further development of epidemiological strategies to contain COVID-19 pandemic and also in the clinical context.

## Data Availability

The data is available with the corresponding author and can be provided on valid request.

## Ethics approval and consent to participate

The study was cleared by institutional ethical committee.

## Acknowledgment

The authors gratefully acknowledge all the healthcare workers for their tireless dedication at each level to fight COVID-19. The authors are thankful to Indian Council of Medical Research, New Delhi for providing financial grants for this study.

---

**Figure 1.**
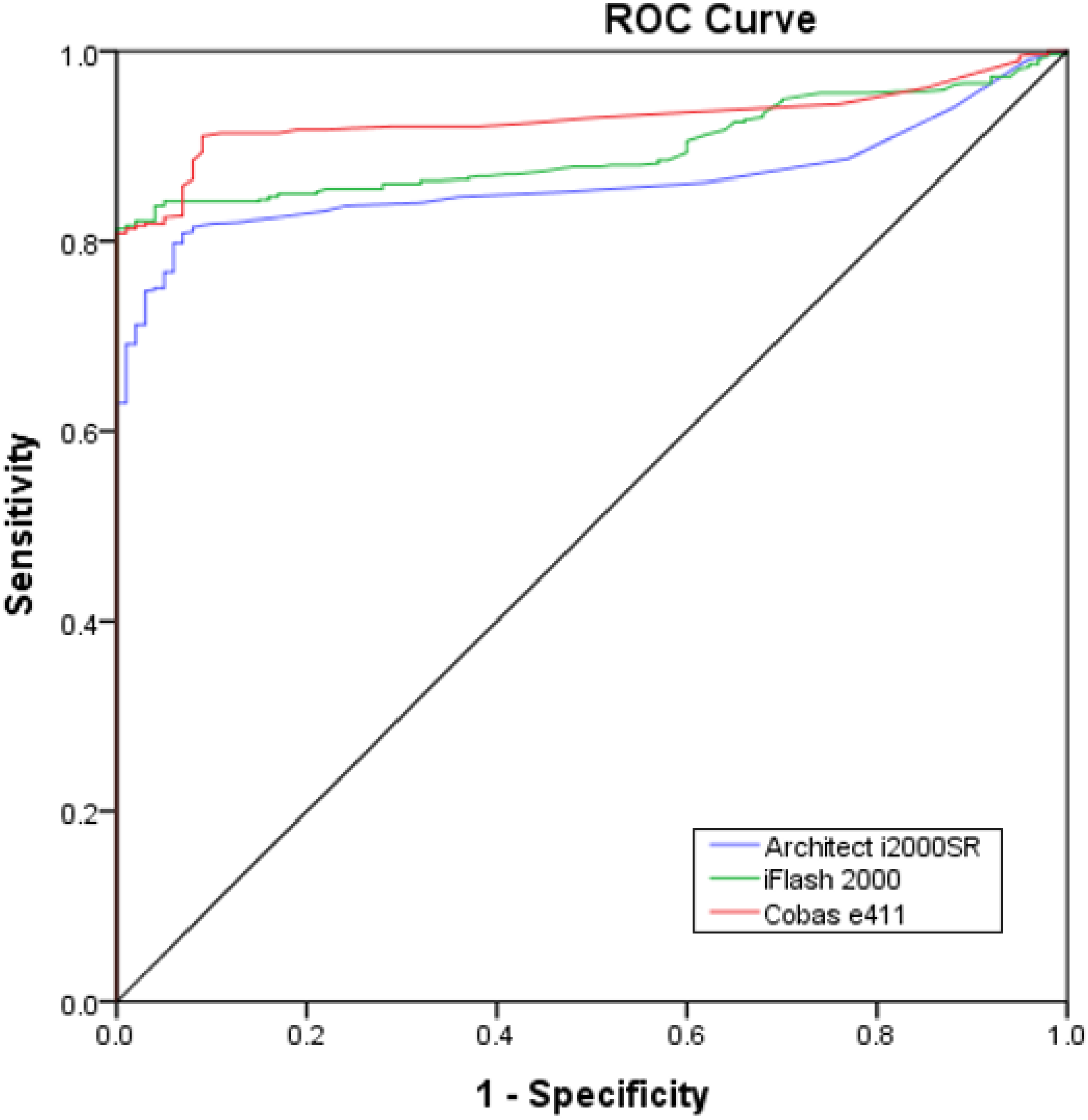
Comparison of diagnostic accuracy by receiver operating characteristic (ROC) curves for Architect i200SR, Cobas e411 and iFlash 1800.

